# Facilitated transition in HIV drug trial closure: a conceptual model for HIV post-trial care

**DOI:** 10.1101/2020.11.14.20231712

**Authors:** Sylivia Nalubega, Karen Cox, Henry Mugerwa, Catrin Evans

## Abstract

Within the HIV clinical trial field, there are gaps in existing ethical regulations in relation to post-trial care. There is need to develop post-trial care guidelines that are flexible and sensitive to local contexts and to the specific needs of different groups of participants, particularly in low income contexts. Evidence regarding HIV trial closure and post-trial care is required to underpin the development of appropriate policies in this area. This article reports research from Uganda that develops a new model of ‘Facilitated Transition’ to conceptualize the transition process of HIV positive trial participants from ‘research’ to ‘usual care’ health facilities after trial conclusion. This was a qualitative grounded theory study that included 21 adult HIV positive post-trial participants and 22 research staff, undertaken between October 2014 and August 2015. The findings showed that trial closure is a complex process for HIV positive participants which includes three phases: the pre-closure, trial-closure, and post-trial phases. The model highlights a range of different needs of research participants and suggests specific and person-centred interventions that can be delivered at different phases with the aim of improving health outcomes and experiences for trial participants in low income settings during trial closure. Further research needs to be done to verify the model in other contexts and for other conditions.

## 1. INTRODUCTION

During the closure of an HIV clinical trial, existing ethical guidelines stipulate a range of obligations for researchers. These include a need to ensure continued access to required HIV treatments and other services,(1, 2) hence measures are required to link trial participants back to adequate services (‘usual care’) once the trial has concluded. Good practice also includes the provision of trial feedback to participants.(3, 4) There is very little evidence available however to guide current practice in this area or to determine the adequacy and appropriateness of existing guidance, particularly in low income settings.(5, 6)

Several authors have criticised existing trial guidance for its emphasis on generic and universalist principles that provide little detail on how to adapt and apply such principles into highly diverse local contexts.(6-8) There is a call to develop post-trial care guidelines that are flexible and sensitive to local contexts and to the specific needs of different groups of participants. In line with good research practice and with the principles of patient and public involvement,(9) such guidance should be developed collaboratively by a range of different stakeholders such as the government, researchers, communities, and research sponsors,(10, 11) and implemented in research practice.(12)

In low income settings such as Uganda, there is often a disparity between healthcare provided in facilities associated with clinical research trials and that provided within general healthcare facilities.(8) Health care provided to participants during the research period varies among individual trials and is guided by regulations set out by regulatory authorities such as the Research Ethics Committees, the sponsors, and the communities involved. The specific nature of care also depends on the characteristics of particular trials (i.e. the likely risks involved, the type of intervention, and the bargaining power of the local authorities). At the minimum, the standard of care in a research facility should not be lower than that provided in public facilities.(13)

In low income settings such as Uganda, it is a reality that most aspects of healthcare provided in clinical trial contexts (in research clinics) is usually significantly better than public sector provision.(7) It usually includes prompt, adequate and free medical treatment for various illnesses for research participants (and sometimes their families), comprehensive medical check-ups, and a range of financial and material incentives such as assistance with transport and meals.(5) Indeed, studies have shown that the superior nature of research-facility based health services can act as a key motivation for individuals to participate in clinical trials,(14, 15) and their withdrawal at trial closure can result in feelings of loss and other negative effects.(16, 17) Hence it is paramount to consider trial closure as an important part of the trial and for this to be managed appropriately. To date, there is no research on the post-trial experience in relation to HIV care in low income settings.(14)

This article presents a model that can guide the post-trial care process, based on findings from a research study that explored how care is perceived and enacted in HIV drug trial closure in Uganda.(18) The model, entitled ‘Facilitated Transition’ generates new insights for researchers and other stakeholders on how to plan and provide holistic and person centred care during closure of clinical trials involving HIV positive participants in a low income setting.

## 2. RESEARCH METHODOLOGY

The research aimed to establish an in-depth understanding of the post-trial care phenomenon amongst participants in HIV drug trials in Uganda. It adopted an interpretive-constructivist grounded theory approach.(19) The study included participants from three trials. Trials 1 and 3 were conducted by the same research institution but at different sites. Trial 1 was conducted at an urban site, situated in Kampala the capital city of Uganda, while Trial 3 was conducted at a peri-urban site, in the Eastern part of Uganda. Trial 2 was conducted at a peri-urban site in the Western part of Uganda. Table 1 below provides a summary of the included trials’ characteristics.

**Table 1:**
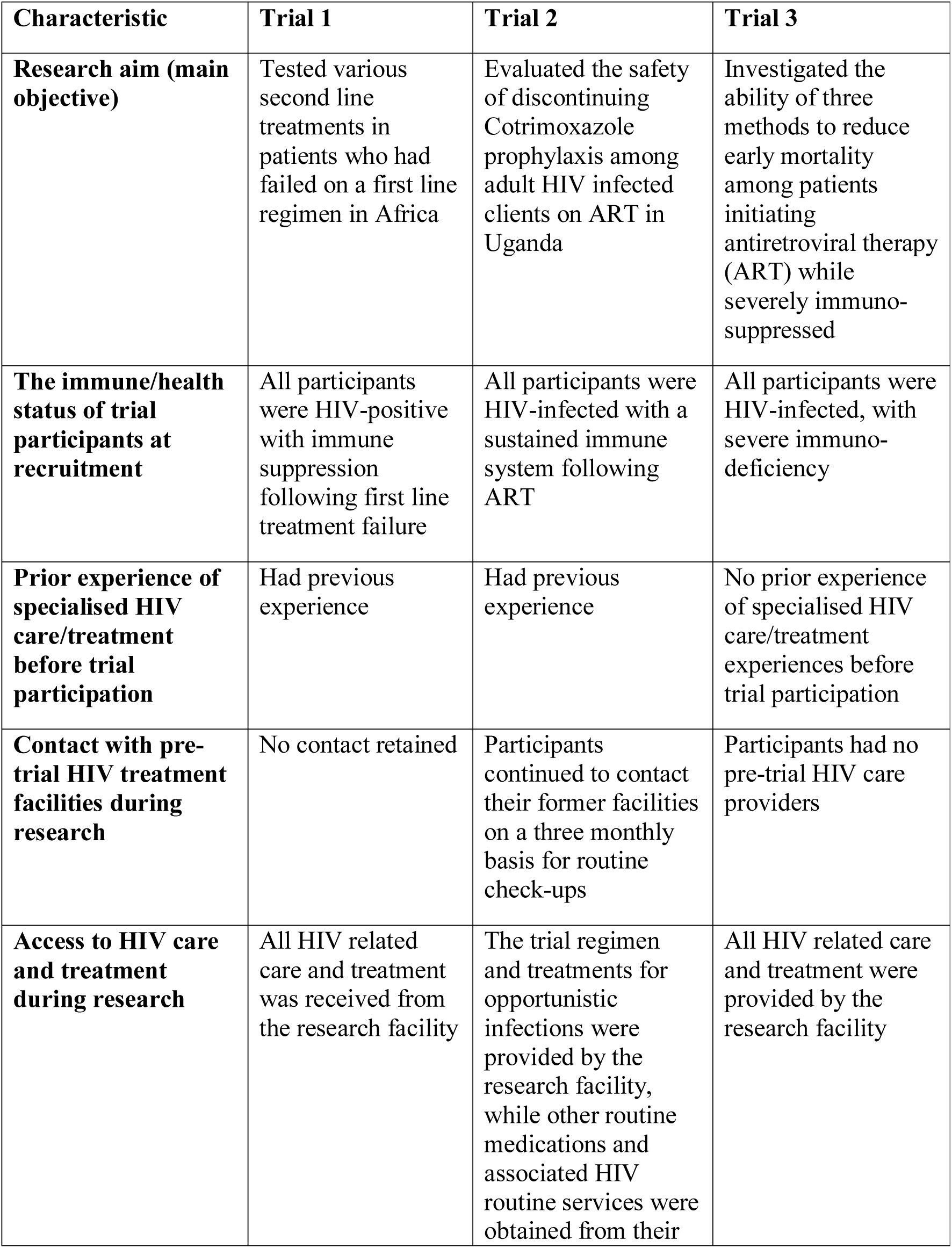
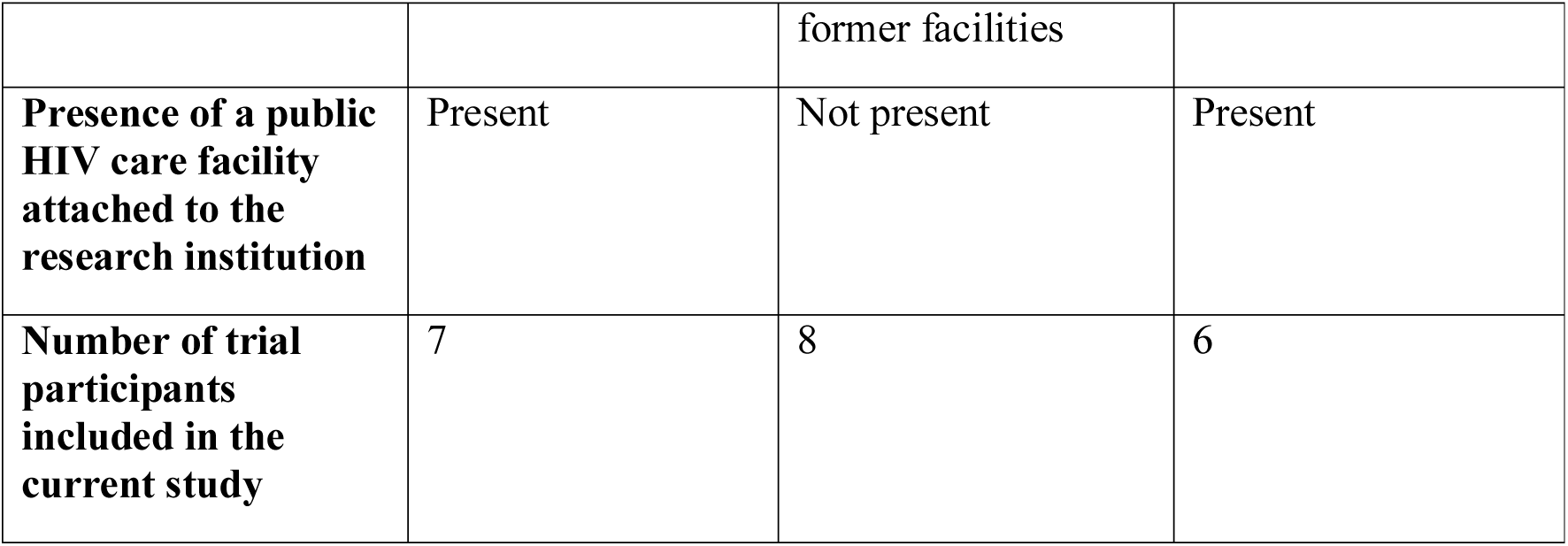
Characteristics of included trials.

| **Characteristic** | **Trial 1** | **Trial 2** | **Trial 3** |
| --- | --- | --- | --- |
| **Research aim (main objective)** | Tested various second line treatments in patients who had failed on a first line regimen in Africa | Evaluated the safety of discontinuing Cotrimoxazole prophylaxis among adult HIV infected clients on ART in Uganda | Investigated the ability of three methods to reduce early mortality among patients initiating antiretroviral therapy (ART) while severely immuno-suppressed |
| **The immune/health status of trial participants at recruitment** | All participants were HIV-positive with immune suppression following first line treatment failure | All participants were HIV-infected with a sustained immune system following ART | All participants were HIV-infected, with severe immuno-deficiency |
| **Prior experience of specialised HIV care/treatment before trial participation** | Had previous experience | Had previous experience | No prior experience of specialised HIV care/treatment experiences before trial participation |
| **Contact with pre-trial HIV treatment facilities during research** | No contact retained | Participants continued to contact their former facilities on a three monthly basis for routine check-ups | Participants had no pre-trial HIV care providers |
| **Access to HIV care and treatment during research** | All HIV related care and treatment was received from the research facility | The trial regimen and treatments for opportunistic infections were provided by the research facility, while other routine medications and associated HIV routine services were obtained from their former facilities | All HIV related care and treatment were provided by the research facility |
| **Presence of a public HIV care facility attached to the research institution** | Present | Not present | Present |
| **Number of trial participants included in the current study** | 7 | 8 | 6 |

### 2.1 Recruitment and data collection methods

The sample strategy aimed to achieve a balance between geographical sites and genders and was guided by the principle of theoretical saturation. Purposive, convenience, and theoretical sampling approaches were employed in selecting participants. The study included 21 post-trial participants and 22 research staff. Confidentiality was ensured by having all eligible trial participants contacted by their former research institutions, to seek their permission to be contacted by the researcher. Those who consented were then approached using phone calls or home visits by the first author. Research staff were approached directly within their institutions, through their supervisors. All but three participants who were contacted agreed to participate in the research. The eligibility criteria for the study is presented in Table 2 below.

**Table 2:** Eligibility criteria.

| **Considered aspects** | **Eligibility criteria** |
| --- | --- |
| Trials | Involved HIV positive participants<br>Involved an HIV treatment or prevention drug<br>Lasted for at least two years<br>Was within four to 12 months of trial closure |
| Post-trial participants | HIV positive<br>Participated in a prevention or treatment drug trial<br>Participated in the trial for at least six months<br>Within three to twelve months of trial exit |
| Research staff | Worked directly with trial participants<br>Was involved in trial closure processes within the past one year |

Data from post-trial participants was collected using in-depth interviews, while a mixture of Focus Group Discussions (FGDs) and Key Informant Interviews (KIIs) were used to collect data from research staff. Participants were asked open ended questions about their experiences of trial closure. All interviews were audio recorded and field notes were taken by the lead researcher to capture additional information of relevance. Data collection followed the constructivist Grounded Theory approach by Charmaz, where initial data analysis informed later data collection.(20, 21) The first author, who was a PhD student at the time of the research, administered all interviews. This author was fluent in the two languages (Luganda and English) in which the interviews were collected, hence there was no requirement to have translators. All interviews were undertaken in the research clinics which the respective trial participants had attended. Participants were compensated with refreshments and a sum of Ugx10,000 (approx. £2.5) mainly to contribute to their transport refund. Data was collected during October 2014 to August 2015.

### 2.2 Data analysis

Interviews were recorded, transcribed, and, where necessary, translated before analysis by the first author. NVivo 10 software was used to organize and manage the data. Data analysis followed a standard grounded theory approach.(19, 21) An initial line by line coding process enabled an openness for any possible theoretical interpretations from the data. Some initial codes were conceptual in nature and were later upgraded to theoretical categories. We then undertook focused coding to elicit more conceptual codes that were representative of the data. This process eventually led to the identification of concepts that were further transformed into tentative categories. Further analytical techniques involved memo writing, theoretical sampling, constant comparison, and diagramming. These analytical techniques led the team to identify a core category ‘Facilitated Transition’,(19, 22) that was developed, through further analysis, into the Facilitated Transition Model.

### 2.3 Maintaining rigor

Attention to rigor was achieved through on-going discussions within the research team regarding each step of the research process and the resultant interpretations from the data.(23) In addition, we strove to enhance credibility by using verbatim quotes to support our interpretations of the data and paying attention to disconfirming cases and opposing or divergent views of the participants. Credibility of the main findings was also enhanced as a result of extensive triangulation of the data (from different sites, different trials and different participant groups).

### 2.4 Reflexivity

We took note of possible influences that the researchers’ theoretical and practice perspectives could have on the research. The first author had previously worked as a research nurse in Uganda. This had exposed her to some of the concerns that HIV post-trial participants face during trial closure. Hence, this background had potential to influence her approach to the research, and her interpretations of the data. We tried to minimize this possible influence through constant discussions with other team members and careful memoing, a process that constantly challenged her pre-suppositions.

### 2.5 Ethical approval

Ethical approval was received from the University of Nottingham, UK and The AIDS Support Organization (TASO) Uganda, Research Ethics Committee (REC). The study was registered with the Uganda National Council for Science and Technology (UNCST), as SS3608. Permission to conduct the research was granted by the respective research institutions, and written informed consent was received from all respondents.

## 3. FINDINGS

This study included a total of 43 respondents from three HIV clinical trials. Twenty-one of these were post-trial participants while 22 were research staff. Of the included trial participants, seven were from Trial 1, eight were from Trial 2, while six were from Trial 3. The majority of trial participants (62%) were female. Trial participants were in the age range of 26-59 years, with the majority (67%) being 40 years and above. Only one participant had attained a university degree, and the majority (67%) were below college level, having either stopped at ordinary or primary levels, or had no education at all. Very few trial participants (14%) were in official employment, while the rest depended on small scale jobs, subsistence farming, or other sources of income such as support from families or friends. Ninety percent (90%) of the trial participants resided in rural or peri-urban settings.

Of the research staff, three were trial coordinators, four were clinicians, five undertook counselling and home visiting, and 10 were nurses (one of the trial coordinators was also a nurse). Trial 1 included 15 research staff, Trial 2 included four research staff, and Trial 3 included three research staff. One quarter of the research staff were male. We also reviewed ethical documents from two trials (Trials 1 and 2), which included: the general trial protocols, participant information documents, informed consent documents, and the trial closure ‘Standard Operating Procedure’ (SOP) documents, also sometimes referred to as the Close out ‘Manual of Operations’ (MOP).

### 3.1 The facilitated transition model: understanding trial closure as a process

Below, we describe the main category that was interpreted from the research – the ‘Facilitated Transition Model’. The model is presented as a whole, followed by a more detailed description of its distinct phases and how they inter-relate, supported by participant quotes and contextual narrative.

Our findings revealed that the transition of HIV positive participants from trial healthcare to non-trial (‘usual’) care facilities was a complex and multi-faceted process that occurred over time and with specific phases. This ‘transition process’ encompassed the events which occurred when an HIV positive trial participant, following planned completion of trial participation, was exited from research and linked back to the public healthcare system, to continue accessing HIV services. One trial participant referred to this transition as “*moving between two worlds*”.

The main events which occurred along the transition process related to trial participants’ care expectations, needs, experiences, and decisions, and to the actions of researchers intended to facilitate and support them. Our research showed that trial participants moved through 3 phases of the trial closure experience during the transition process, which include: (i) the pre-closure phase (which represents events or experiences that occurred before the actual trial closure but that influenced the post-trial care experiences), (ii) the trial closure phase (which is the active phase of the closure in which trial participants were prepared and exited from the trials), and, (iii) the post-trial phase (which represents events that occurred after trial participants had been exited from trials until 12 months later). These phases are demarcated by time points that reflect how the transition process evolved, proceeded and concluded. Our findings also identified facilitative actions and strategies that were or could be implemented at each stage to support the transition process. These include: (i) planning for post-trial care (in phase 1), (ii) providing emotional and practical support (in phase 2) and (iii) providing psycho-social support (in phase 3). The process and its phases are depicted in Figure 1, in a model of ‘Facilitated Transition’, followed by a description of each phase of the model in turn.

**Figure 1:**
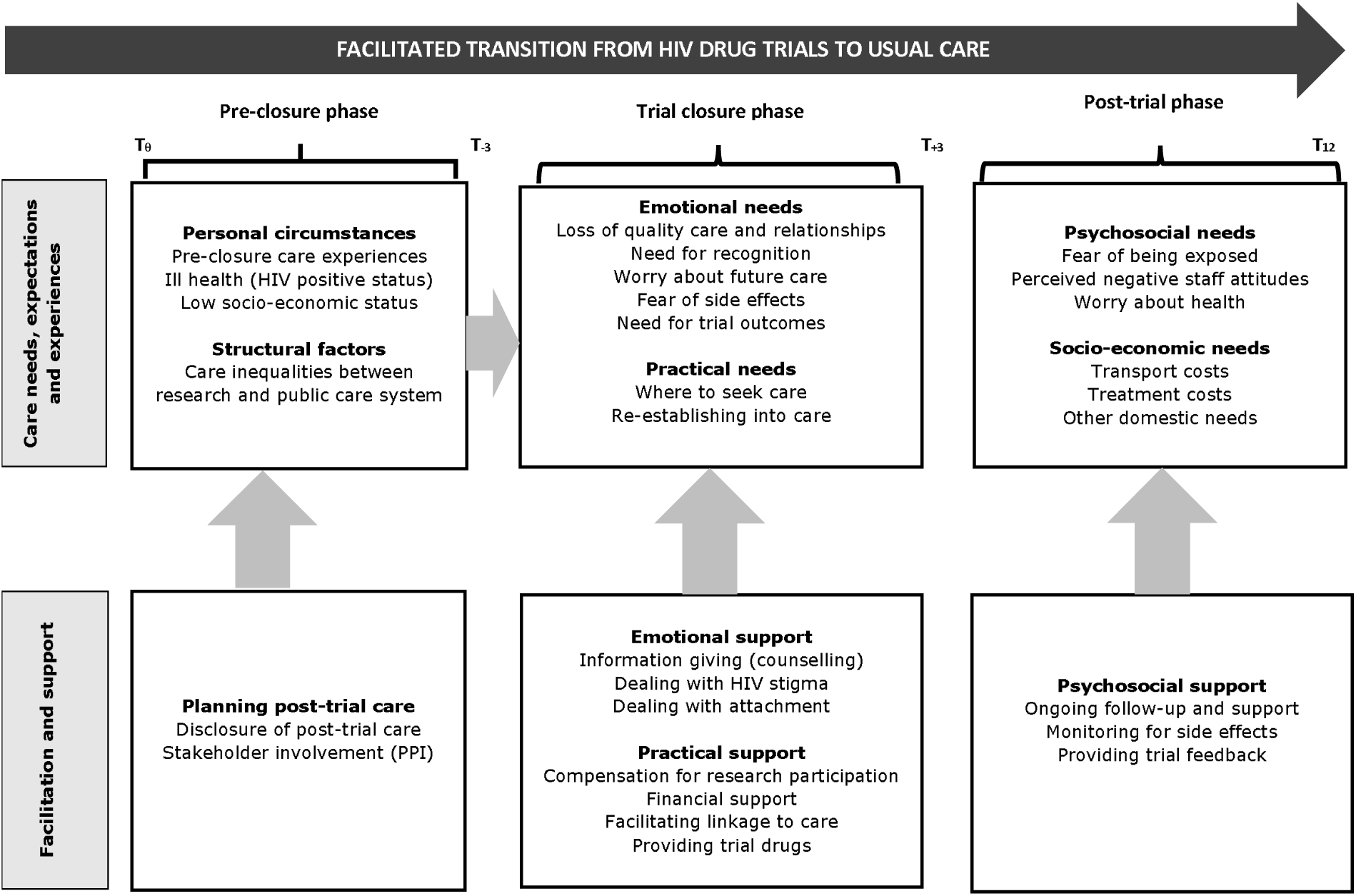
Model of Facilitated Transition.

### 3.2 Pre-closure phase

According to the study participants, the pre-closure phase involved activities undertaken or required during the preparation of participants for trial closure. This phase was guided by policies on post-trial care that clarify the minimum post-trial care expectations from researchers and is underpinned by trial participants’ care needs, expectations and experiences. The pre-closure phase starts before or during planning of the trials up to three months before the actual exit of the participant from the trial **(T**_θ_ **-T**_**-3**_).

> *Trial closure starts at the beginning of the study apparently. Because as we start the treatment of patients, we go through the screening, we go through the enrolment, we see them settle in, we prepare them or we tell them that there is time when the study will end so that as we start, as they settle in, they know that there is time and the trial will end. So, by the time we get to the closure, they are already into closure. (Joy, counsellor/health visitor/community mobiliser)*

Several contextual factors that existed prior to trial participation were found to influence how participants reacted to the closure and how they made decisions about post-trial care. These factors include those related to (i) individual participants and (ii) those related to the Ugandan health care structures.

Personal factors such as ongoing ill health and poverty, and participants’ experiences at the end of previous trials in which they may have participated influenced their care needs, expectations and experiences during the closure of the current trial. For example, participants with ongoing health issues (such as opportunistic infections) during trial closure were worried about how they would receive care after they left research, and these worries were exacerbated for those living in poverty. In addition, some participants had previous experience of being involved in research, and their previous experiences influenced their expectations of post-trial care in the current trial (for example if they had previously been given money or other benefits):

> *… because I will refer so much to the other one (trial) …… while exiting us, they also gave us some money. Yes and now this one, they just exited us, and only gave us transport. So now that is why I have told you that I will insist on that, to show you that we didn’t receive any money as such, yet in the other one people were many in the FX study because there was good money. But this one had no money as such. (Baker, Trial 2)*

Participants’ perceptions and experiences related to previous healthcare delivery were also found to influence how participants reacted to trial closure. In the current study, a strongly held perception of ‘poor’ quality of care in the Ugandan public healthcare facilities caused fear and tension amongst trial participants as they reflected on where they would seek care after the trials ended. Hence, inequalities in care provision between research facilities and the public health care facilities in Uganda was a contributing factor to negative emotions among trial participants during trial closure.

> *When we were brought from there (the former care sites), my understanding was that they have removed me from that place because it was no longer capable. So when you get me from there and you bring me here and you send me back after giving me some treatment, and you send me back there, I think I cannot be contented. Why, because you got me from there and brought me, now again you have completed the research and you are taking me back, it means that I may go back to the same situation as I was. (Joel, Trial 1)*

During this pre-closure phase a key factor thought to facilitate post-trial care was early discussion about post-trial care plans (where trial participants were informed about the trial closure processes from the start of the trial). Discussions of when the closure will occur, how participants will be exited from the research, where they will be linked for post-trial care, how trial drugs will be accessed after the trial, and when/how trial results will be disseminated, were made. This activity helped to psychologically prepare the participants for trial closure.

> *As research teams, one of the obligations we have is to ensure that before research participants or trial volunteers are exited, they are prepared. One, when you start, your recruitment must be actually, it is in the consent form that they know the period they are going to be in the study. So once they know that may be three years, they should be working towards three years. (Bernard, community liaisons officer)*
>
> *In the beginning when they were informing us about the research, they told us that it (the research) would last for three years, after we have started, and when you have spent three years, for you…, so when they ended, they exited us. (Abdu, Trial 2)*

Policy guidelines were a useful tool for guiding researchers on post-trial care preparation. Ethical documents such as study protocols, informed consent forms, and trial closure/exit documents were among the documents that contained information that guided trial closure processes.

> *Transferring patients back into standard clinical care: A list of participants with dates of exit will be sent to individual referral/mother centres prior to exit. Sites will be informed that participants will be supplied with drugs enough for 3 months at exit and that their next appointments will be on individual appointment’s card at exit. Each participant will be given an exit report to the referral centre but this will be followed by a telephone call to confirm that the patient delivered the report and that the appointment date was noted and recorded in their appointments management system for follow-up. (MOP, Trial 1)*
>
> *A clinical summary report will also be the referral form. (This) will be produced in triplicate by the data manager at each site. One copy ---kept by the participant, one ---taken to the ART provider (referral letter) by the participant, one kept in the participant’s file. (MOP, Trial 2)*

### 3.3 Trial closure phase

The trial-closure phase describes events that occurred around the point of trial closure. It included active trial closure activities such as psychological preparation, exiting, and linking trial participants to post-trial care facilities. This phase began three months prior to the planned exit of a participant from the clinical trial until three months after the exit **(T**_**-3**_**-T**_**+3**_). Research participants reported that the intensive preparation for trial closure started on average three months to the planned closure, a period which marked off the trial closure phase as reflected on the Facilitated Transition model.

> *The period (of preparing for trial closure) is inbuilt in our community engagement process, so I don’t want to say three months before the trial ends or six months. It is inbuilt. […] So our post-trial engagement does not start say, when we have seen the last volunteer, its inbuilt we share it, but the bridging activities that take us between when people are seen last for their trial participation and when the results are out, and what happens beyond the results are out is what I actually I would say begins three months before the end of the trial. (Bernard, community liaisons officer)*

In the trial-closure phase, trial participants were found to have a range of emotional and practical support needs. Participants experienced multiple emotions resulting from the trial closure and concerns about how to access care afterwards. The main factors that influenced participants’ emotional reactions included: the loss of quality care and valued relationships in research settings, the need for recognition (compensation) for their contributions in research, worry about future care after leaving research, fear of side effects from trial interventions, and the need to receive feedback on trial outcomes.

> *What I felt as for me was that we were going to miss the treatment we were receiving from here. Because at health facility Y, sometimes there is no medications for treating other illnesses you may suffer from and so you have to buy that medication, yet here we would get it. (Abdu, Trial 2)*

The worries about access to post-trial care were generally associated with the perceived poor healthcare in the Ugandan public healthcare facilities. Hence, participants reported concerns about how best to find alternative care facilities and how to link to them. Many participants had left their previous ‘usual’ care facilities when they joined the clinical trials while others had no HIV care experience prior to taking part in the trials. These participants were now required to identify suitable healthcare facilities for continuation of HIV care and treatment. The good care experiences in the research centres coupled with perceptions about, and experiences of, poor care in the public healthcare facilities influenced how trial participants made choices of where to seek post-trial care.

> *The thing which made me to select this place (site C), they handled me 99, let me say 100%. The way they handled me is not like other hospitals, that is why I selected to stay here. (Nandi, Trial 3)*
>
> *The care I got there (in research), it was just enough. I thought I am not going back to that place (pre-trial care facility), I have to go back to site A. Yes, the care and ok the distance also, but the care most. (Janet, Trial 1)*

Where new facilities were identified, participants also cited challenges in reporting to them, in terms of the complex procedures required for registering into a new facility. They reported this process as inconvenient and difficult and expressed a desire to be supported by the research team, to be assisted to register.

> *So in order to make it easy for us, they would do the same thing. They would get a person with their referrals and take him/her back to facility D and they open for you another file. So you are not disturbed… (Sumin, Trial 1)*

To facilitate the trial closure phase, researchers engaged in offering a range of emotional and practical support strategies to address the care needs of the participants.

> *Then of course, all the time we have to talk to the patients because some we know become a bit anxious, they have been with you for four years, may be for how many years, now somehow the end is coming, so you have to keep preparing them. (Jane, trial coordinator)*
>
> *So the linkage is providing them with a referral form with the details of their clinical picture which they deliver to the service provider and then it gets filed in their records the other side. (Ivan, trial coordinator)*

### 3.4 Post-trial phase

The post-trial phase describes events which occurred after a trial participant has been exited from the trial and linked to a new facility where they would continue to access HIV care. This phase starts approximately three months following trial closure and continues up to 12 months later **(T**_**+3**_**-T**_**12**_).

> *…. there is that extra follow-up that may be done probably at 90 days, but continually it poses a challenge especially when finances have to be incurred. (Elhana, nurse)*
>
> *Actually, if it were possible, it is post-trial, it would be better that may be you know what is happening after a year. You know for a year at least you know… (Jane, trial coordinator)*

The post-trial phase illuminates how HIV positive trial participants adapted to the post-trial situation, whilst receiving HIV care and treatment within the Ugandan public healthcare system. Although trial documents were generally silent on most aspects of this phase, researchers and trial participants recognised the need for supporting HIV positive post-trial participants during this phase. A number of psychosocial and socio-economic factors influenced trial participants’ care needs, expectations and experiences during the post-trial phase.

The psychosocial concerns that arose following participants’ exit from HIV trials included fear of HIV stigma (since non-trial care facilities did not provide as much privacy as research institutions) and fear of poor healthcare (including overcrowding in the facilities, lack of privacy, and poor time management), and specific concerns about how their individual healthcare needs would be met (such as accessing routine HIV medications and treatments for opportunistic infections). Trial participants also reported negative staff attitudes (e.g. rudeness) which affected clinic attendances.

> *And I went and informed them ‘I am having headache,’ but it was not a clinic day, and I informed them I was having headache but they never gave me any medications, not even Panadol, in fact I went away crying. (Brenda, Trial 2)*

In addition, some participants were concerned about the possibility of side effects resulting from trial interventions, and wished to be informed of any plans of follow up and monitoring them after leaving the trials.

> *Because the other side (post-trial care facility), it is just a matter of presenting your…, may be your book and they write down and they send you to the pharmacy and they pick up your medication, but they cannot know the changes which is taking place within you. So the first people (researchers) they are the ones who are good whereby if you have any problem you can call them and you can go and they try to examine you again and see what is going on. (Wilberforce, Trial 3)*

In terms of socio-economic issues, participants reported experiencing significant challenges in meeting the financial burden of accessing care (for example transport and treatment costs), after leaving the trials. Those with ongoing ill health problems were affected more significantly by the transition as these were unlikely to be working yet suddenly required additional finances (e.g. to buy medications or had specific food requirements).

> *…we have to look for money for transport to fetch medication. And when I fail to get it, I call them (research staff) and I request them to help me with money to go and I collect the medication. (Ruth, Trial 1)*

Participants in the current study reported a number of measures that are required to support trial participants during the post-trial phase, to enable them to cope with both their psychosocial and economic needs. The supportive measures included; establishing mechanisms to ensure and check that post-trial participants had successfully linked to alternative care facilities and are receiving appropriate treatments, providing continued psychological support, material and financial support, and monitoring for possible side effects. Providing such support required continued engagement of research staff and other stakeholders with the participants during the post-trial phase.

> *… what they would have done is to continue moving, to continue inquiring to know after they have gone back to where they were (before joining research), how have they experienced life where they are; […] I would say that this is the real summing up (concluding) of the research. (Baker, Trial 2)*
>
> *I believe we are supposed to be following them up, to see how they are coping up, to see the challenges they are facing, and like these Trial 3 patients that we recruited with low CD4s, to find out may be if they have increased, they are rising up, to see generally how they are improving. (Favour, Counsellor/home visitor)*

However, findings indicated that in current practice, there was very little engagement of researchers in the post-trial phase, despite this being recognised as a very important part of trial care.

> *It is really very important though we don’t actively do it, post follow-up. Because when they go back to their mother clinics, a lot of things change, it is like transitioning. So if you are transitioning, you need to be followed up until you settle in properly. (Charlotte, counsellor/home visitor)*

In general, the lack of post-trial care was blamed on inadequate policy guidance for researchers in terms of how to support this phase of the trial. It was felt that the existing guidelines lacked detail on important aspects of post-trial care which became a challenge for researchers to plan for and implement them.

> *And then, the other thing we have to put into mind is when you are budgeting, you have budgeted for the study up to the exit, up to the closure. So you don’t have more funds to cater for people after this time, may be even the staff have been employed up to that time. All these things keep, really tie us. […] So even if you wanted, really you can’t. (Destiny, nurse)*

The participants recommended that policy makers include post-trial care into their guidelines to make it mandatory for the researchers to provide post-trial support.

> *So I would recommend that it is put into policy that every trial conducted, especially clinical trials conducted, they should do a post-trial assessment to know how their patients are doing. (Alloy, trial coordinator/nurse)*
>
> *If it is an obligation or if it is a policy of an institution, then they can add it (post-trial follow-up) on the budget; it can be added onto the budget and say ‘for us we do this, if it is a policy of an institution. (Jane, trial coordinator)*

## 4. DISCUSSION

The model presented in this paper shows that clinical trial closure is a complex process for HIV positive participants in Uganda, and can be conceptualised as having 3 different phases, each with a range of emotional, socio-economic and practical needs requiring different support strategies from trial staff. The model is supported by a recent study (albeit in a different context) that similarly reported emotional and psychosocial concerns as negative trial-closure outcomes. (7) In line with Cho and others,(1) our findings emphasise the importance of providing care and support to trial participants to ensure a smooth transition from research to non-research (‘usual’) care.

In this study, both research staff and participants expressed a strong need for a more facilitated approach to the trial closure process, in terms of supporting participants to make the transition back to usual care facilities at the end of a trial and maintaining contact to ensure queries around side effects and infromation around trial outcomes could continue to be shared. The ‘Faciltated Transition’ model describes key facilitation/care strategies that could be applied to support participants during each of the three phases. The care needs of post-trial participants varied depending on individual circumstances such as health status, economic status, and available supportive structures, thereby requiring a case by case approach to post-trial care, while trying to observe the general guidelines. Hence, the Facilitated Transition Model depicts a person centred approach, and considers the need for ethical practice within a flexible context-based framework.(24)

The pre-trial phase sets the pace for post-trial care by identifying trial participants’ expectations of post-trial care and is the beginning of preparation for trial closure. It is recommended that post-trial care activities and outcomes (such as potential harms) be disclosed to the participants before consent to participate, to facilitate the informed consent process.(7) Relevant policies play a role at the pre-trial phase, to guide researchers on what is expected of them in regards to post-trial care.(25, 26) The guidelines are expected to be comprehensive enough to include activities that will be done to meet the post-trial needs of the participants.(6)

The trial-closure phase highlights the main fears and concerns of trial participants associated with leaving trial-related healthcare and finding suitable alternatives. The psychological concerns that arise within the trial-closure phase mainly relate to the loss of the quality care(8) in research related facilities which cannot be matched with that in the public sector.(7) A recent systematic review similarly identified the loss of research-related care as a cause of stress among post-trial participants and that affects their integration into the public services.(16) Post-trial participants also go through a financial burden associated with continued access to treatments especially for opportunistic infections (and especially among those with low immunity). Various interventions are required to support the trial participants during this phase, including psychological, financial and material support.(27, 28) In addition, participants found a challenge of locating and reporting to new service providers, and suggested that the process of linking to care needed to a more active approach from the trial researchers.(29)

The post-trial phase is the period where trial participants adjust and get integrated into the public healthcare system following trial exit. Participants report unique needs during this phase that may affect their health and wellbeing, including access to HIV treatment and prevention drugs, having to deal with the routines of the public healthcare system which are largely unfavourable,(30) and looking for financial wellbeing. Since linkage to HIV care is not a one off event,(30) our study reccomends the need for supporting trial particpants during the post-trial phase, through a follow up mechnism and continous engagement by research staff (and other stakeholders), to enable them to get well established into the ‘new’ healthcare system.(1)

A crosscutting concern across the whole transition journey is the lack of guidelines in support of post-trial care. It was reported that limited support was provided in the post-trial phase, as most activities were not actively planned for by the researchers. The trial documents that were reviewed as part of the study found that they lacked guidance on some of the important post-trial care aspects, as observed by other researchers.(6, 12) Our study reccomends the need for policies on post-trial care to be strengthened, by highlighting the important post-trial care requirements and enforcing them in research practice.

### 4.1 Transferability of the conceptual model

The model of Facilitated Transition is primarily intended to guide the practice of closure of HIV drug trials invoving HIV postive individuals in Uganda, by providing flexible guidelines which can be applied to different HIV drug trial contexts and geographical settings in Uganda. These guidelines can however be applicable in other situations such as HIV drug trial closure in other low income settings, transitioing of post-trial participants with other chronic illnesses such as cancer, transitioning of individuals hospitalised for long periods and re-establishing into the community/other healthcare settings, and settling of other groups of individuals in the community e.g. from prisons.

### 4.2 Limitations

This was a single qualitative study that included a relatively small sample of participants, which limits generalizability of the research findings beyond the studied sample and context. Additionally, this study presented a potential for recall bias given that it was undertaken retrospectively with some participants being interviewed close to 12 months following trial exit. A larger prospective study on a similar population would improve understanding of the studied phenomenon.

### 4.3 Conclusions

This article expands on the understanding of trial closure and post-trial care in HIV drug trials involving HIV positive individuals in low income settings, by describing trial closure as a process rather than a one-off event. The Facilitated Transition model explains the process of transitioning from HIV research to ‘usual care’ facilities and offers guidelines on a person-centred approach to post-trial care. By illustrating the concerns and needs of the participants, and suggesting possible support approaches to address these, the model can thus be useful in the planning and provision of post-trial care for HIV positive trial participants in Uganda and related settings. This approach is likely to result into favourable health outcomes for the participants.

## Data Availability

All data referred to in this manuscript can be accessed with permission from the corresponding author.

## 5. ACKNOWLEDGEMENTS

We acknowledge the participants who participated in this study for their time and valuable views, the participating institutions and their stuff for their effort and support during study conduct, and the funders for SN’s PhD study (Vice-Chancellor’s Scholarship for Research Excellence (International) and the University of Nottingham, United Kingdom) under which the current study was undertaken.

## Notes

### Competing Interest Statement

The authors have declared no competing interest.

### Funding Statement

This research received no specific grant from any funding agency in the public, commercial, or not-for-profit sectors. The Ph.D programme under which the study was undertaken was sponsored by the Vice-Chancellors Scholarship for Research Excellence (International) and the University of Nottingham, United Kingdom.

